# Effects of silver diamine fluoride on oral bacteriome and mycobiome: a randomized clinical trial

**DOI:** 10.1101/2024.03.11.24304114

**Authors:** Mayura Manerkar, Vivianne Cruz de Jesus, Betty-Anne Mittermuller, Victor H. K. Lee, Sarbjeet Singh, Mary Bertone, Prashen Chelikani, Robert J. Schroth

**Affiliations:** Rady Faculty of Health Sciences, University of Manitoba, Winnipeg, Manitoba, Canada; Children’s Hospital Research Institute of Manitoba, Winnipeg, Manitoba, Canada; Shared Health Inc., Winnipeg, Manitoba, Canada

**Keywords:** dental caries, microbiome, bacteria, fungi, children, Clinical Trials, Randomized

## Abstract

**Introduction:** Silver diamine fluoride (SDF) is a simple and non-invasive agent used to arrest early childhood caries (ECC). This study aimed to investigate potential changes to the oral microbiome in children with ECC who were treated with SDF at three different frequency regimens.

**Methods:** Forty-five children (*n*=15 per group) with ECC were recruited into a randomized clinical trial testing three different treatment frequency regimens of SDF. A total of 195 carious lesions were treated with two applications of 38% SDF and 5% sodium fluoride varnish (NaFV) and assessed over three study visits (one month (Regimen 1M), four months (Regimen 4M), or six months (Regimen 6M) apart). Dental plaque samples were collected at each visit. Sequencing of the V4-*16S rRNA* and *ITS1 rRNA* genes were used to study the supragingival plaque microbiome.

**Results:** The overall arrest rates for treated carious lesions were 75.9% at Visit 2 and 92.8% at Visit 3. Arrest rates were higher for all lesions after two applications of SDF with NaFV, and applications one month and four months apart had higher arrest rates (95.9% and 98.5%) than six months (81.1%) apart. The microbial diversity analyses showed no significant differences in the overall microbiome after SDF treatment. However, significant changes in the abundance of specific bacteria and fungi, particularly *Lactobacillus spp.*, *Bifidobacterium spp., and Candida spp.* were observed after treatment. Furthermore, overabundance of *Streptococcus mutans* and *Candida dubliniensis* at baseline was observed in children who had at least one caries lesion not arrested after one SDF application, compared to those who had 100% arrest rates.

**Conclusion:** SDF with NaFV applications were an effective modality for arresting ECC, with higher arrest rates after two SDF applications. No loss of diversity but significant changes in the abundance of specific bacteria and fungi were consequences of SDF treatment.

## Introduction

Early childhood caries (ECC), characterized by the presence of tooth decay in primary teeth in children younger than 6 years of age, is a multifactorial disease caused by a combination of factors including oral colonization with increased levels of cariogenic bacteria, decreased microbial diversity, and teeth that are susceptible due to enamel defects (e.g., hypoplasia) and/or frequent consumption of fermentable carbohydrates [1–3]. Within Canada, day surgery for ECC constitutes 31% of all day surgeries for children aged 1-5 years, making this the major cause for day surgeries within this age group [4, 5].

*In vitro* studies have shown that interkingdom interactions were associated with dysbiosis of the oral microbiome [6, 7]. Both *Candida albicans* and *Streptococus mutans* develop symbiotic interactions in the presence of sucrose that boost their ability to colonize teeth and synergistically enhance virulence which results in aggressive onset of dental caries lesions [6].

Treatment of young children with severe ECC often requires high-cost general anesthesia and advanced clinical skills which may be unavailable or unaffordable for many pediatric populations [8]. Oral rehabilitation may involve multiple extractions, restorations and crowns [9]. Children treated under general anesthesia often develop new or recurrent caries within months of surgical rehabilitation, with some children requiring subsequent visits to the operating room for dental treatment [10, 11]. Traditional treatment modalities do not address causative factors and have not reduced the incidence of childhood caries and rates for dental surgery in North America [5].

Silver diamine fluoride (SDF) is a simple and non-invasive topical fluoride solution to arrest caries in children. A solution of 38% SDF (Advantage Arrest, Oral Science, Brossard, QC, Canada) contains 25% silver, 5.5% fluoride, and 8% ammonia. In the pilot study that preceded our current study, Sihra et al. (2020) concluded that at least two applications of SDF are recommended, with lesion arrest rates of 74.1% and 96.2% after 1 and 2 applications of SDF, respectively [12]. Application of SDF in children too young to tolerate restorative procedures can stabilize active caries until the child is older and able to cooperate for traditional restorative techniques or for minimally invasive techniques [13]. Since treatment with SDF is non-invasive and easily performed [14], it can be a useful addition to existing traditional treatment modalities. Primary teeth that have been treated with SDF and atraumatic techniques can sustain function and act as a space maintainer until eruption of the permanent successor [14].

Previous studies characterized the effect of SDF treatment on the plaque bacteriome [15, 16] and mechanistic studies investigated the effect of SDF on only few *Candida spp.* [17, 18]. However, the influence of the application regimen of SDF on the oral mycobiome is not yet studied. The purpose of this study is to investigate the oral bacteriome and mycobiome changes that result from SDF applications randomized to three different application frequency regimens. This study is part of a larger randomized clinical trial to investigate the effectiveness of using SDF to arrest ECC in children.

## Methods

### Study Population, Design, and Sample Collection

The study protocol was approved by the University of Manitoba Biomedical Research Ethics Board (HS24849-B2021:031) and was registered on ClinicalTrials.gov (NCT04054635). Study visits took place at the Children’s Hospital Research Institute of Manitoba (**CHRIM**) or at community-based dental clinics in Winnipeg (Access Downtown, Mount Carmel Clinic and SMILE plus). A total of forty-five children < 72 months of age were recruited based on inclusion and exclusion criteria (**Supplementary Table 1**) between December 2019 and June 2020 with block randomization by site in order to achieve equal proportions in each regimen by clinic site of recruitment.

All children had ECC and were randomized to three different treatment regimens (*n* = 15 per group; **Supplementary Table 2**), in an open-label parallel trial design, using sealed envelopes containing a regimen number. The sealed envelopes were generated by the research coordinator. When a child was recruited, a research personnel randomly selected an envelope, assigning the child to one of the groups. Each treatment protocol was assigned a regimen number (1, 2 and 3) for randomization purposes. Regimen 1 was the Winnipeg Regional Health Authority (**WRHA**) Oral Health Program Protocol, advising two applications of SDF four months apart. Our pilot study by Sihra et al. (2020), used the WRHA protocol so this regimen was chosen as Regimen 1 [12]. Regimen 2 was the protocol suggested by the American Dental Association (**ADA**), recommending two applications of SDF six months apart [19]. Regimen 3 was the protocol proposed in the American Academy of Pediatric Dentistry (**AAPD**) Clinical Practice Guideline suggested by Horst, recommending an initial application followed by 2-4 weeks of monitoring, with reapplication if needed [20–22]. We implemented this as two applications of SDF one month apart. For simplicity, Regimen 1 (4 months), Regimen 2 (6 months) and Regimen 3 (1 month) will henceforth be referred to as Regimen 4M, Regimen 6M and Regimen 1M respectively.

Written informed consent, by parents or caregivers, was obtained for each child. Dental exams, assessment of lesions, supragingival plaque collection, and SDF applications were completed by a dentist. Carious lesions were assessed based on clinical hardness (very soft, medium, or very hard), colour change (yellow, brown, or black), and size of lesions. The colour, hardness, size, and the dmft (decayed, missing, and filled primary teeth) scores were recorded for each visit. Hardness was assessed with light pressure within the lesion using a probe. Teeth were also assessed for pain and/or infection at each visit. Lesions were deemed arrested if found to be black and very hard.

Supragingival plaque samples were taken by scrubbing a sterile interdental brush on all available tooth surfaces [23, 24]. Samples were then dislodged into 1 mL of RNAprotect Reagent (Qiagen, Hilden, Germany) and then frozen at −80°C until DNA extraction. Dental plaque samples were taken at first (baseline), second, and third (final) study visits. Silver Diamine Fluoride 38% (Advantage Arrest, Oral Science, Canada) was applied to cavitated lesions involving dentin with a microbrush for one minute. The surface was then wiped with wet gauze or rinsed with water. SDF application was followed by application of 5% sodium fluoride varnish (NaFV; NuPro White, Dentsply Sirona, USA).

Demographic data, arrest rates, and dmft scores were analyzed using NCSS 2022 statistical software (NCSS, LLC). The data were analyzed using Anova, Kruskal-Wallis, Chi-square, and paired sample t-test, when appropriate. *P* < 0.05 was considered significant.

### DNA Extraction, 16S and ITS1 rRNA Amplicon Sequencing and Data Analysis

DNA extraction was completed using the DNeasy PowerSoil Pro Kit (Qiagen, Hilden, Germany). The plaque samples were centrifuged for 10 minutes at 13,000 rpm. The supernatant was then discarded and 800 µL of buffer CD1 was used to re-suspend the resultant pellet. This mix was then added to the PowerBead Pro Tube and manufacturer’s instructions were followed for DNA extractions.

The library preparation and paired-end Illumina MiSeq PE300 sequencing of the V4 region of bacterial *16S rRNA* and fungal *ITS1* (Internal Transcribed Spacer 1) *rRNA* genes were performed by Genome Quebec Innovation Center. The forward and reverse primers used to amplify the *16S* and *ITS1 rRNA* genes, respectively, were the V4-16S 515F (GTGCCAGCMGCCGCGGTAA) and 806R (GGACTACHVGGGTWTCTAAT), and the ITS1-30F (GTCCCTGCCCTTTGTACACA) and ITS1-217R (TTTCGCTGCGTTCTTCATCG) [25].

### Bioinformatics and Statistical Analysis

Sequencing data was analyzed using QIIME2 2018.11 (Quantitative Insights into Microbial Ecology) [24, 26]. DADA2 implemented in QIIME2 was used to filter and merge the pair-end sequences, and to obtain the table of amplicon sequence variants (ASVs) [27]. The ITS1 sequences were trimmed using the Q2-ITSxpress QIIME2 plugin before the DADA2 step [28]. The Human Oral Microbiome Database (HOMD, version 15.1) and the UNITE database (version 8.2; QIIME developer release) were used for the taxonomic assignment of bacteria and fungi, respectively, with classify-sklearn naïve Bayes taxonomy classifier in QIIME2 [23, 29–31]. The fungal ASVs that were assigned only at kingdom level were submitted to further fungal ASV curation with the R package LULU [32] and the program BLASTN in NCBI [33]. The ASVs with non-fungal BLASTN results were discarded and the remaining were repeatedly assigned to new taxonomic assignments using different UNITE databases threshold levels [31, 34, 35] and taxonomy classification methods (q2-feature-classifier classify-sklearn and classify-consensus-blast) in QIIME2 [24, 36]. The data was imported into R using the R package “qiime2R” (version 0.99.13) and additional filtering was performed using “phyloseq” (version 1.30.0) to remove singletons [37, 38]. The ASV counts were then normalized using the cumulative-sum scaling (CSS) approach from the R package “metagenomeSeq” version 1.28.2 [39].

The alpha diversity analysis was performed using the Shannon index and raw ASV count data in R (“phyloseq” package, version 1.30.0). Alpha diversity comparisons were done by the Kruskal-Wallis test or Friedman test as appropriate. Beta diversity analysis was performed on CSS normalized ASV data, using weighted UniFrac distances and the permutational analysis of variance (PERMANOVA) test with 999 permutations in the R package “vegan” (adonis function; version 2.5.6) [40]. It was visualized using principle coordinate analysis (PCoA) in the R package “ggplot2” (version 3.3.3) [41].

A paired DESeq2 negative binomial Wald test was used to detect differentially abundant species between groups, controlling the false discovery rate (FDR) for multiple comparison [42]. FDR adjusted *p* < 0.05 was considered significant.

## Results

### Characteristics of Study Participants

Forty-five children, 15 children per group, with mean age of 43.5 ± 13.8 months participated. Seventeen (37.8%) were female and 28 (62.2%) were male (**Table 1**). One child assigned to Regimen 6M did not return for follow-up, bringing final sample size to 44 participants and 132 plaque samples. No harm or unintended effect was observed.

**Table 1.** Characteristics of study participants.

| Age at Baseline (months)<br>Mean ± SD | | | *p*-value |
| --- | --- | --- | --- |
| Total | 43.5 ± 13.8 | | |
| Regimen 1M | 43.7 ± 14.15 | | 0.09 |
| Regimen 4M | 41.6 ± 12.7 | | |
| Regimen 6M | 45.2 ± 15.1 | | |
| Sex, n (%) | Female | Male |  |
| Total | 17 (37.8%) | 28 (62.2%) |  |
| Regimen 1M | 4 (26.7%) | 11 (73.3%) | 0.51 |
| Regimen 4M | 7 (46.7%) | 8 (53.3%) |  |
| Regimen 6M | 6 (40%) | 9 (60%) |  |

### Lesion-Level Analysis

A total of 195 carious lesions in 44 children were treated at baseline and followed over two subsequent study visits. One child presented with an abscessed SDF-treated tooth at Visit 3 in Regimen 1M and this lesion was considered a failure i.e., not arrested. Average arrest rates were higher at Visit 2 and Visit 3 for Regimen 4M and Regimen 1M compared to Regimen 6M (**Table 2**). Arrest rates were higher for all lesions after two applications of SDF, with a statistically significant increase in the arrest rate from Visit 2 to Visit 3 for Regimens 4M (*P* = 0.015) and 1M (*p* < 0.001) (**Table 2**).

**Table 2.** Average dmft score and arrest rates.

|  | Average dmft |  |  | Average Arrest Rate |  |
| --- | --- | --- | --- | --- | --- |
|  | Visit 1 | Visit 2 | Visit 3 | Visit 2 | Visit 3 |
| Regimen 1M | 6.67 | 6.73 | 6.73 | 81.1% (60/74) | 95.9% (71/74) |
| Regimen 4M | 6.40 | 6.40 | 6.40 | 80.9% (55/68) | 98.5% (67/68) |
| Regimen 6M | 6.79 | 6.93 | 6.93 | 62.3% (33/53) | 81.1% (43/53) |
| **Total** | **6.61** | **6.68** | **6.68** | **75.9%<sup>a</sup>**<br>**(148/195)** | **92.8%<sup>b</sup>**<br>**(181/195)** |

dmft: decayed, missing, and filled primary teeth.
<sup>a</sup> There was a significant difference between Regimens 1M, 4M, and 6M at visit 2 (Pearson's Chi-square test, $p < 0.05$ ).
<sup>b</sup> There was a significant difference between Regimens 1M, 4M, and 6M at visit 3 (Pearson's Chi-square test, $p < 0.05$ ).

Average dmft score remained the same between Visit 1, 2, and 3 for Regimen 4M. While for Regimens 1M and 6M, the average dmft increased between Visit 1 and 2 and remained the same at Visit 3 (**Table 2**).

### Supragingival Plaque Bacterial Community Analysis

The alpha diversity analysis showed no significant difference between visits for all regimens, although a slight increase in the average diversity can be observed between visits 2 and 3 (Friedman Test, *p* > 0.05; **Fig. 1A**). Overall, the beta diversity analysis showed no significant differences in the supragingival plaque bacteriome between visits (**Fig. 1B**).

**Figure 1.**
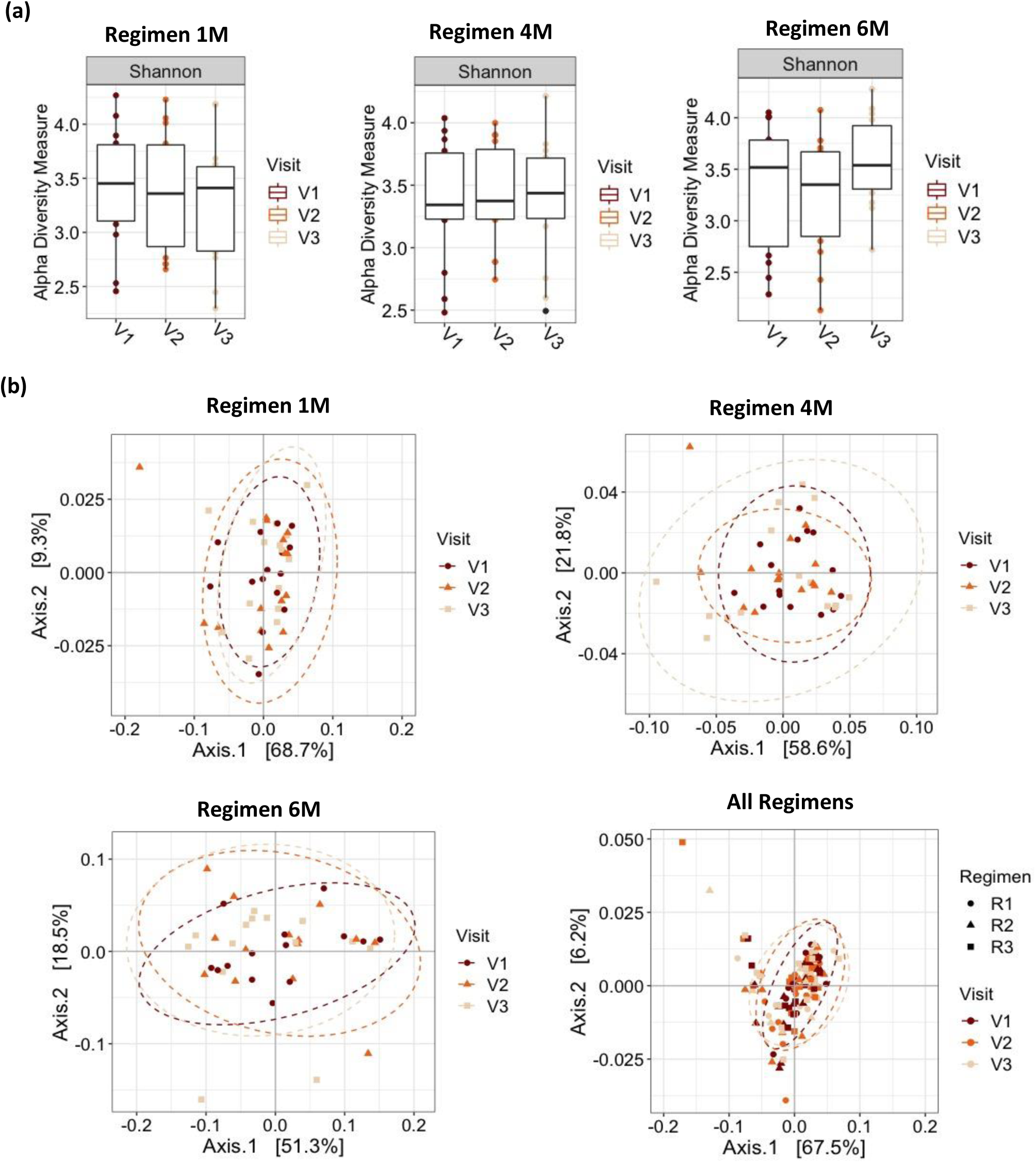
Alpha and beta diversity analyses. (a) Alpha diversity analysis. Boxplot of Shannon Index for bacterial taxa in Regimen 1M, Regimen 4M and Regimen 6M and subgroup analysis by visit. The line inside the box represents the median. Whiskers represent the lowest and highest values within the 1.5 interquartile range. (b) Beta diversity analysis. Principal coordinate analysis plots of weighted UniFrac distances based on the overall structure of the supragingival plaque bacteriome. Each data point represents a sample, coloured according to visit. The ellipses represent a 95% confidence level. No significant separation of samples was observed between regimens or visits.

Taxonomic assignment showed that *Streptococcus*, *Corynebacterium* and *Actinomyces* were the most abundant genera overall (**Fig. 2A**). Remarkably, *Streptococcus mutans* showed a decrease in relative abundance at Visit 3 compared to Visit 1 for Regimen 4M and Regimen 1M. For Regimen 6M*, S. mutans* showed an increase in relative abundance from Visit 1 to Visit 2 and then a decrease in relative abundance from Visit 2 to Visit 3 (**Fig. 2B**). However, these changes were not found to be statistically significant.

**Figure 2.**
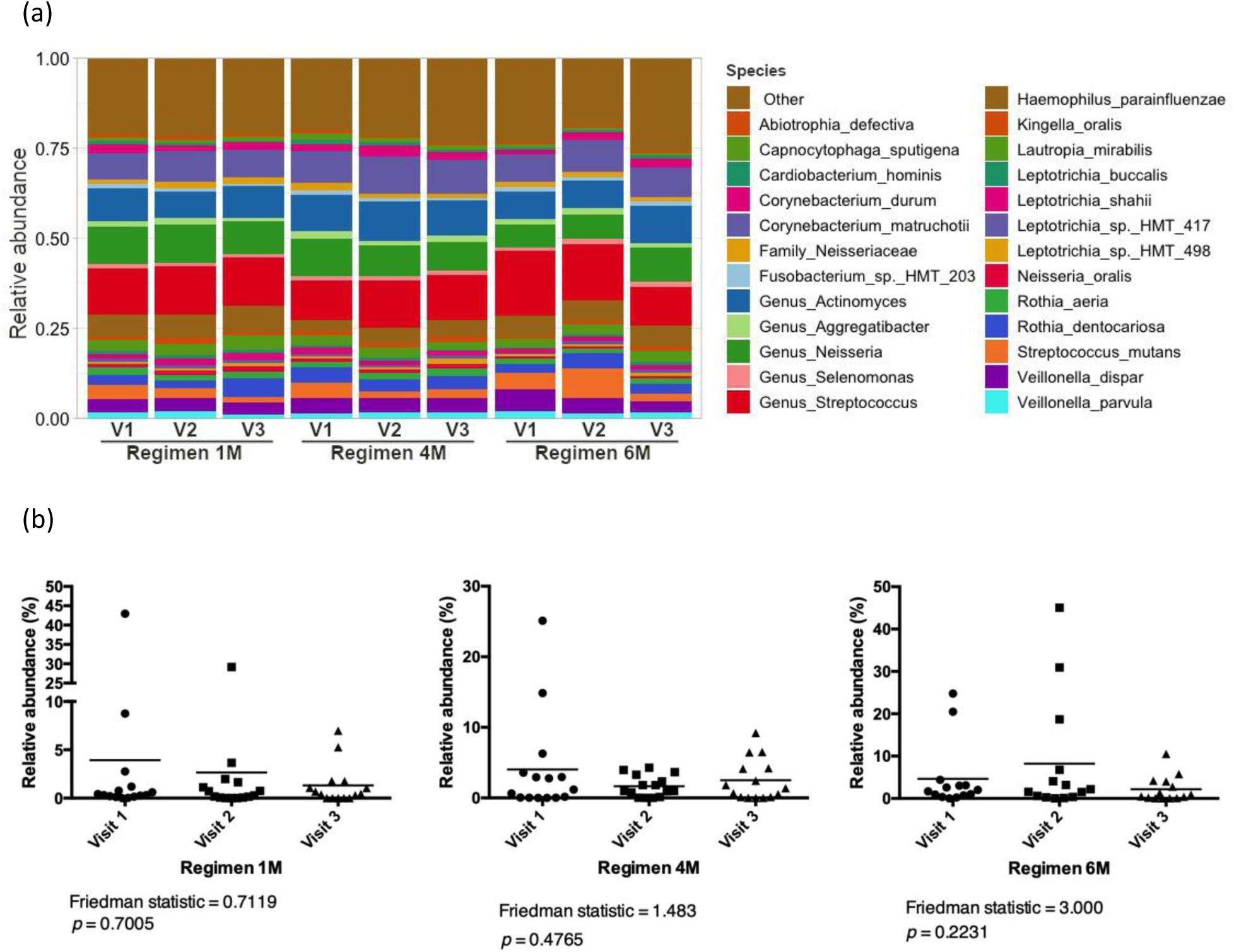
Bacterial taxonomic profile of dental plaque and relative abundance of *Streptococcus mutans*. (a) Taxonomic profiles of dental plaque according to regimen and visit, at bacterial species level. Colours were assigned only to the top 25 most abundant taxa. (b) Relative abundance of *S. mutans* according to regimen and visit.

The differential abundance analysis showed that various bacterial species were significantly enriched or depleted between Visits 1 and 2 or between Visits 1 and 3 (**Fig. 3** and Supplementary Figure 1-4). Interestingly, compared to the following visits, the supragingival plaque of the children at baseline (Visit 1) was enriched with cariogenic bacteria such as *Lactobacillus spp.* and/or *Bifidobacterium spp.,* regardless of the regimens (Supplementary Figures 1-4). However, children in Regimen 6M showed higher abundances of *Lactobacillus salivarius* in Visits 2 and 3 and also higher dmft scores at Visits 2 and 3 and lower arrest rates, compared to the other regimens (Supplementary Figure 3, **Table 2**).

**Figure 3.**
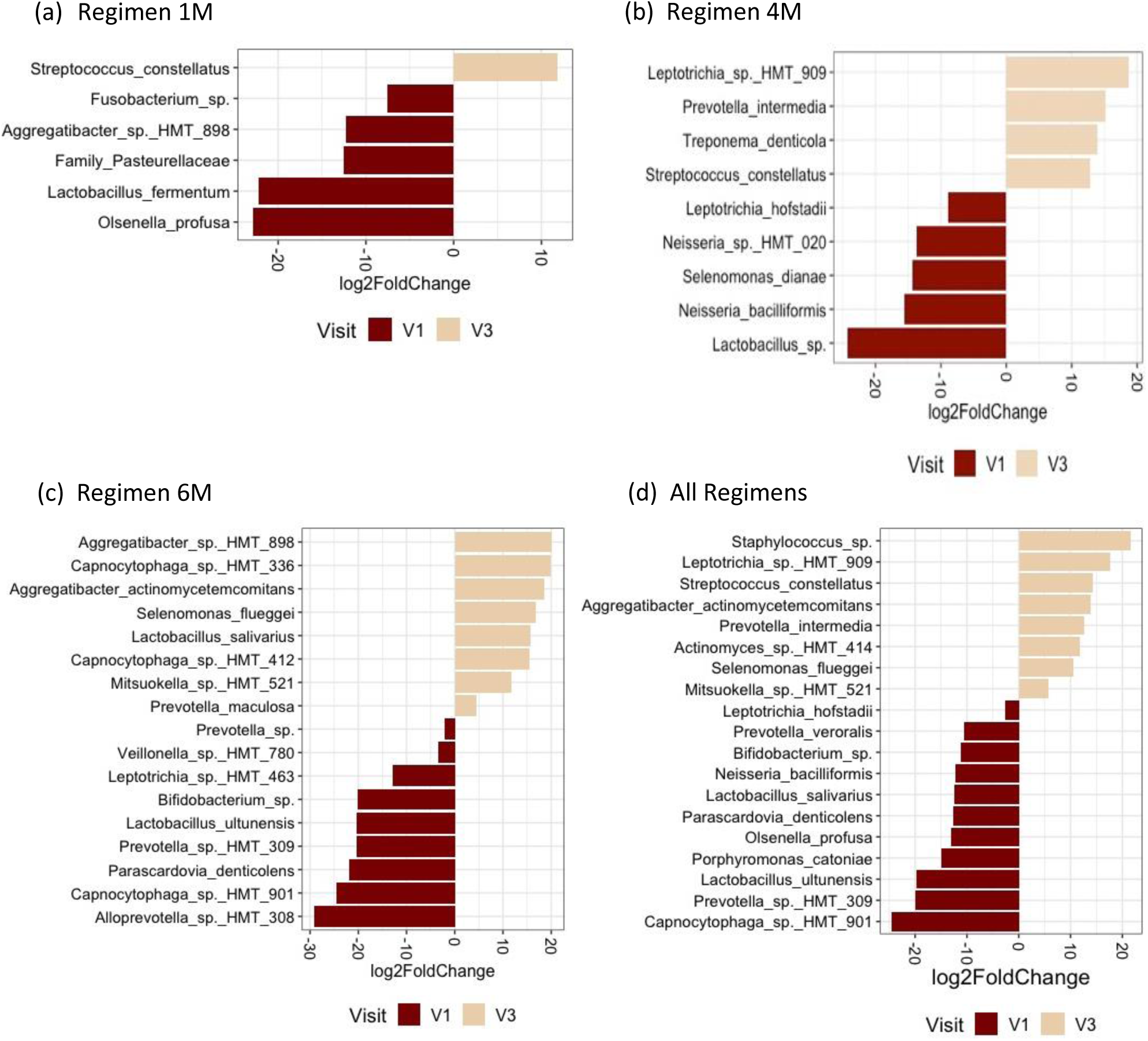
Differential abundance analysis for bacterial species comparing visits 1 and 3. The figure shows the relative fold change in bacterial abundance between visits 1 and 3 in (a) Regimen 1M, (b) Regimen 4M, (c) Regimen 6M, and (d) All regimens together. Only bacterial taxa with FDR adjusted *p* < 0.05 are shown.

### Supragingival Plaque Fungal Community Analysis

Five samples did not pass quality control filtering steps for the mycobiome data analysis and, due to the design of the study, a total of eighteen samples were removed to allow for paired analysis of the remaining 114 plaque samples. The alpha diversity analysis showed no significant differences between visits for all groups (Friedman Test, *p* > 0.05; **Fig. 4A**). Similarly, the beta diversity analysis showed no significant difference in supragingival plaque mycobiome between visits for all groups (**Fig. 4B**).

**Figure 4.**
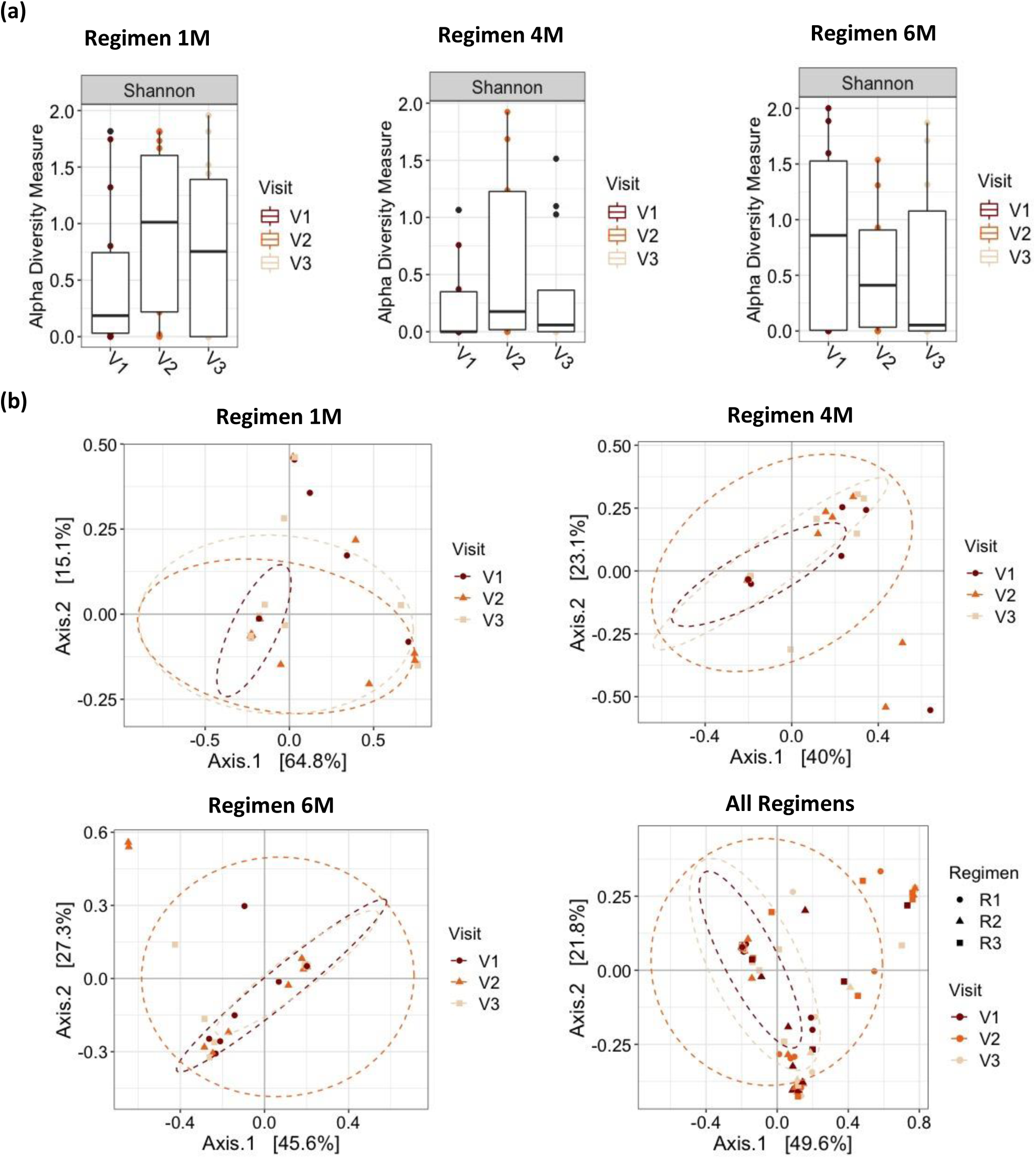
Alpha and beta diversity analyses of fungal microbiome. (a) Alpha diversity analysis. Boxplot of Shannon Index for fungal taxa in Regimen 1M, Regimen 4M and Regimen 6M and subgroup analysis by visit. The line inside the box represents the median. Whiskers represent the lowest and highest values within the 1.5 interquartile range. (b) Beta diversity analysis of fungal communities. Principal coordinate analysis plots of weighted UniFrac distances based on the overall structure of the supragingival plaque mycobiome. Each data point represents a sample, coloured according to visit. The ellipses represent a 95% confidence level. No significant separation of samples was noted between regimens or visits.

Taxonomic assignment showed that *Candida, Blumeria*, and *Malassezia* were the most abundant genera. **Fig. 5** shows the relative abundances of the top 25 most abundant fungal taxa and it shows that *Candida albicans* was highly abundant in all groups regardless of number of visits or regimens.

**Figure 5.**
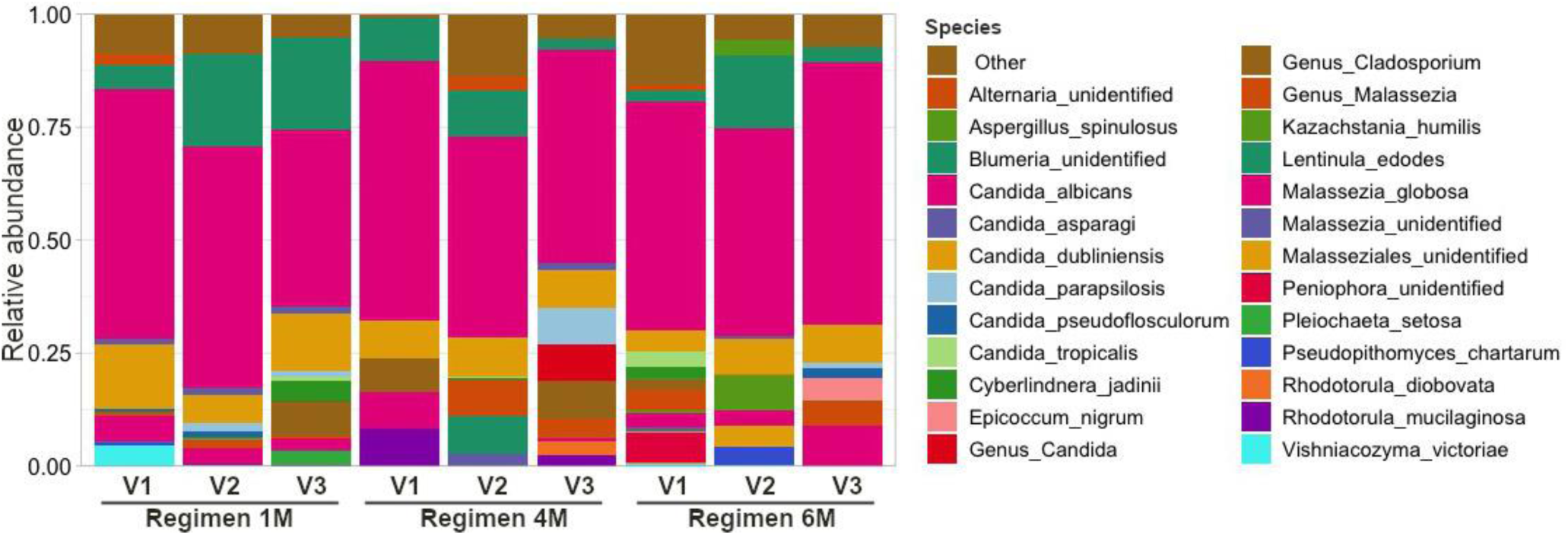
Taxonomic profiles of the children’s dental plaque according to regimen and visit, at fungal species level. Colours were assigned only to the top 25 most abundant taxa.

The differential abundance analysis showed many fungal species that were significantly enriched or depleted between visits in all Regimens (**Fig. 6** and supplementary Figures 2, 3). We noted that there was a significant decrease in *Candida tropicalis* from Visit 1 to Visit 3 in Regimen 6M. This decrease was also seen from Visit 2 to Visit 3 and Visit 1 to Visit 3 when all regimens were taken together. All these results taken together suggest that SDF treatment may have an effect on the abundance of specific fungi but it does not modify the overall microbial structure of the supragingival plaque.

**Figure 6.**
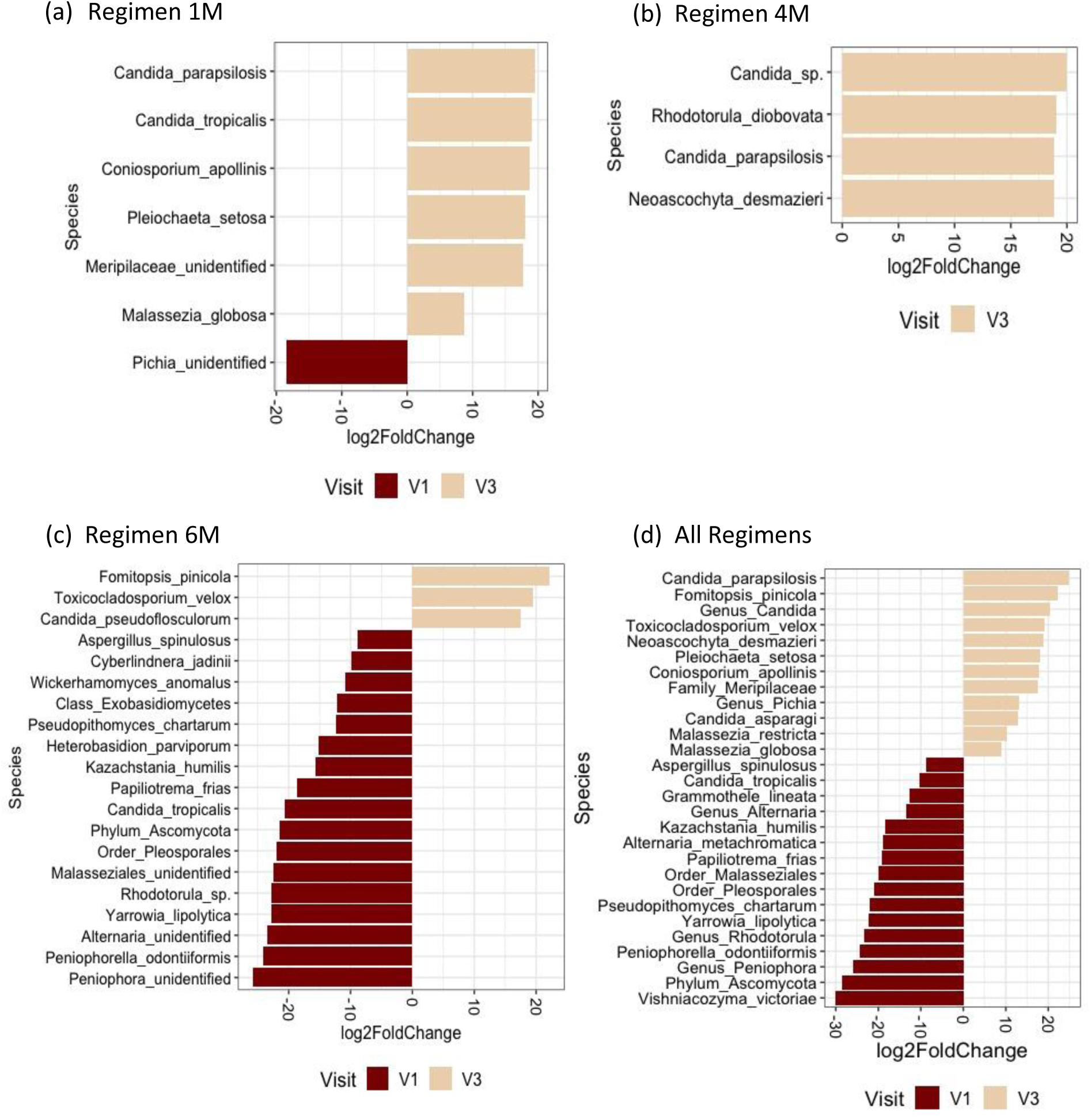
Differential abundance analysis for fungal species comparing visits 1 and 3. The figure shows the relative fold change in fungal abundance between visits 1 and 3 in (a) Regimen 1M, (b) Regimen 4M, (c) Regimen 6M, and (d) All regimens together. Only fungal taxa with FDR adjusted *p* < 0.05 are shown.

### Link between the baseline microbiome and the SDF treatment outcome

After only one application of SDF, 40.9% (*n* = 18/44) of the children had 100% of the treated caries lesions arrested. While, 59.1% (*n* = 26/44) of the children had at least one lesion that was not arrested at Visit 2. Among those children, six had anterior lesions not arrested, 16 had posterior lesions not arrested, and four had both anterior and posterior lesions not arrested. To evaluate if the baseline microbiome is linked to SDF treatment outcomes, we compared the baseline microbiome of children who had 100% of caries arrested after one application of SDF and those with at least one lesion not arrested after one application of SDF (i.e., at Visit 2). For this we used differential abundance analysis adjusting for differences in the regimen used and the sex of the child. Interestingly, the dental plaque of the children with lesions not arrested after one SDF application was enriched with the caries associated species *S. mutans* and *Candida dubliniensis* at baseline, compared with children who had 100% arrest rates (**Fig. 7**).

**Figure 7.**
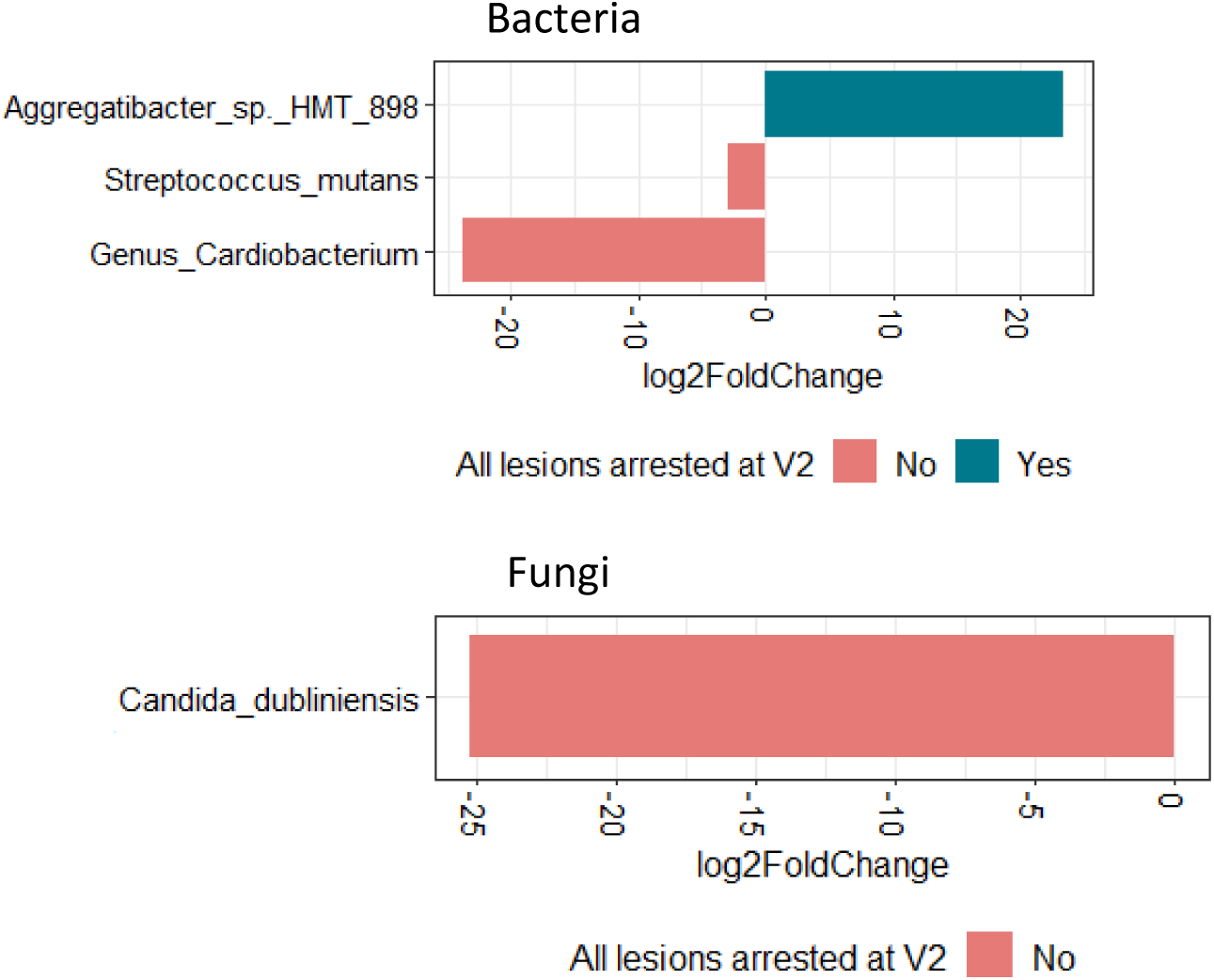
Differential abundance analysis comparing the differences in the baseline microbiome between the children who had 100% of the lesions arrested (Yes) and those with <100% arrest rates (No) after one SDF application. Only baseline (before SDF treatment) microbiome data was used for the analysis.

## Discussion

SDF treatment in children too young to tolerate restorative procedures can stabilize active caries until the child is older and able to cooperate for traditional restorative or minimally invasive techniques [13]. In a pilot study, it was concluded that at least two applications of SDF are recommended, with 96.2% of arrest rate achieved, which is similar to our findings [12]. Since SDF treatment is non-invasive and easily performed, it can be a useful addition to existing traditional treatment modalities [14]. Primary teeth that have been treated with SDF and atraumatic techniques can sustain function and act as a space maintainer until eruption of the permanent successor [14].

### Lesion-Level Analysis

The current AAPD Clinical Practice Guideline recommends monitoring carious lesions for arrest at 2-4 weeks following application of SDF with reapplication as necessary to achieve arrest of all targeted lesions and re-care monitoring based on disease activity and caries risk level at three, four, or six months [22]. SDF is known to have a reservoir effect with sustained antimicrobial effects over time [43]. Application to carious lesions is as effective in preventing caries in other teeth and surfaces as applying SDF directly [43]. In our study, the average dmft remained the same over all three visits for Regimen 4M which is consistent with these findings. Average arrest rates were higher for all groups after two applications of SDF compared to one application of SDF, however, arrest rates were higher with application frequencies of 1 month and 4 months compared to 6 months.

Regimen 6M showed lower arrest rates compared to Regimen 1M and 4M and also showed increased dmft between Visit 1 and Visits 2 and 3. The literature suggests that increasing the frequency of SDF application can increase caries arrest rates [22]. The effectiveness of lesion arrest by 38% SDF decreases over time [22, 44]. Re-activation of treated lesions has been observed in a 2-year study where a single application of 38% SDF was sufficient to prevent only 50% of the arrested surfaces at six months from reverting to active lesions again over 24 months [44]. We suggest that six months between applications may not be optimal for arrest of caries in high-risk children. Other factors which could be influencing arrest rates and dmft with six months application frequency and that need to be investigated in future studies include the number of erupted teeth, diet high in fermentable carbohydrates, plaque index, amount of plaque present in lesions, medical status, and use of antibiotics.

### Microbiome

Previous studies have looked at microbial changes following SDF treatment in pediatric patients [15, 45]. They looked at changes in the bacteriome at the caries lesions only and had smaller sample sizes. Milgrom et al. (2018) found no consistent changes in relative abundance of caries-associated microbes or lesion-specific microbial diversity before treatment and at follow up (14-21 days) for 3 children from each group (38% SDF vs placebo) [45]. In our study, no significant differences in alpha or beta diversity were observed in the overall plaque microbiome after SDF treatment. The lack of loss of diversity following SDF treatment suggests that the use of SDF is safe with minimal risks of growth of opportunistic pathogens [45].

Mei et al. (2020) found no overall microbiome changes in 5-year-old children immediately before, 2 weeks after, and 12 weeks after, one application of 38% SDF [15]. Microbiota showed a temporal shift in the positive direction after two weeks but returned to original status after twelve weeks. The authors noted that *S. mutans* tended to decline in arrested carious lesions while *Neisseria* species and *Actinomyces naeslundii* tended to increase in arrested carious lesions. Carious lesions that remained active after SDF treatment showed a trend of increased abundance of *S. mutans, S. sobrinus*, and *Lactobacillus sp*. compared to pre-SDF treatment levels though not all were statistically significant. Takahashi et al. (2021) have shown that 38% SDF can decrease the number of *S. mutans*, along with the amount of water-insoluble glucan, and the thickness of the formed biofilm *in vitro* [46]. They suggest that this may be partially due to an inhibitory effect of silver ions on glucosyltransferase activity (which is used by *S. mutans* to synthesize extracellular polysaccharides from sucrose) and the rupture of the bacterial cells. This may explain why the supragingival plaque bacteriome showed a decrease in relative abundance from Visit 1 to Visit 3 for *S. mutans* for all regimens, however, this trend was not found to be statistically significant (Fig. 2B).

It was also noted that the plaque microbiome was depleted of other cariogenic bacteria and *Candida* species after two applications of SDF. For instance, when all regimens were taken together, *L. salivarius* was enriched in Visit 1 compared to Visit 2 and Visit 3, signifying a decrease in abundance from Visit 1 to Visit 3. *L. salivarius* is known to be a dominant species commonly isolated from the dentition of adults and children with caries [47].

High levels of *Candida albicans* are frequently detected in the plaque biofilms of children with ECC [48–50]. Other *Candida* species such as *Candida tropicalis*, *Candida krusei* and *Candida glabrata* are also detected in plaque biofilms of children with ECC, though not in the numbers of *C. albicans* [48]. In our previous study on the characterization of the oral microbiome in children with S-ECC, we found that *C. dubliniensis* and *C. tropicalis* were more abundant in oral swab samples of children with S-ECC compared to caries-free controls [24]. In contrast, healthy, ECC-free children have plaque biofilms where *C. albicans* is absent or sporadically detected [48–50]. *Candida albicans* was found to be highly abundant in all groups regardless of the number of visits or regimens. Our differential abundance analysis showed many fungal species that were significantly enriched or depleted between Visits 1 and 2 or between Visits 1 and 3 suggesting that SDF treatment may have an effect on the abundance of specific fungi. A study by Fakhruddin et al. (2020) has shown that SDF has antifungal effects against some Candida species including *C. albicans, C. krusei, C. tropicalis* and *C. glabrata* [17].

### Limitations

Although our sample size was larger than that of previous SDF studies examining the oral microbiome, our sample size was small, and that could be partially attributed to the recruitment of participants during the COVID-19 pandemic and the limited funding for sequencing. Furthermore, although the same number of children were recruited for each group, the total number of teeth treated per group varied, with the Regimen 1 having the highest number of teeth treated (74) and the Regimen 6 having the lowest number of teeth treated (53), due to the loss of one participant in Regimen 6. Furthermore, in this study, plaque samples were not site-specific and so data on the microbiome could not be localized to the site of the carious lesion. Comparing overall microbiome changes with site-specific changes would help determine whether SDF has a more pronounced local effect than overall oral microbiome effect. Most oral fungi are present at low biomass and may be difficult to detect in oral samples [51]. In our study, five samples had low number of sequence reads and a total of eighteen samples had to be removed to allow for paired analysis. As the study focused on ECC and included only children younger than 6 years of age, our findings cannot be generalized for all age groups.

## Conclusions

SDF with NaFV was an effective modality for arresting dental caries with higher arrest rates for all lesions after two applications. Application of SDF 1 month and 4 months apart had higher arrest rates than application 6 months apart suggesting that six months between applications may not be optimal for arrest of caries in high-risk children.

No significant loss of microbial diversity was seen after SDF treatment. Moreover, significant changes in the abundance of bacterial and fungal species, particularly caries-associated species, were identified. This study provides information on the effect of SDF treatment on the overall dental plaque microbiome of young children with ECC. Further studies with a larger sample size are needed to confirm whether the presence or absence of various bacterial and fungal species are the result of SDF applications at various frequencies.

## Data Availability

Raw sequencing data produced in the present study are available upon reasonable request to the authors.

## Acknowledgements

We thank the parents/caregivers, participants, and the staff at the community-based dental clinics in Winnipeg (Access Downtown, Mount Carmel Clinic, and SMILE+).

## Statement of Ethics

### Study approval statement

The study protocol was approved by the University of Manitoba Biomedical Research Ethics Board (HS24849-B2021:031).

### Consent to participate statement

Written informed consent, by parents or caregivers, was obtained for each child.

## Author Contributions

M.M. contributed to data acquisition, data analysis, data interpretation, and writing, critically revised the paper, and approved the final version. V.C.J. contributed to data acquisition, data analysis, data interpretation, and writing, critically revised the paper, and approved the final version. B.M, V.K.L., and S.S. contributed to data acquisition, critically revised the paper, and approved the final version. M.B. and P.C. contributed to study design and data interpretation, critically revised the paper, and approved the final version. R.J.S. conceived the idea for this study, contributed to study design, data acquisition, data interpretation, and writing, critically revised the paper, and approved the final version.

