## Supplementary for "Effects of silver diamine fluoride on oral bacteriome and mycobiome: a randomized clinical trial"

**Supplementary Table 1.** Inclusion and Exclusion Criteria.

| **Inclusion Criteria** | **Exclusion Criteria** |
|---|---|
| **1)** Child is < 72 months of age with ECC, with active carious lesions.<br>**2)** Child has ≥ 1 primary tooth with caries that is eligible to receive SDF.<br>**3)** Eligible primary teeth must have soft cavitated caries lesions extending into dentin [International Caries Detection and Assessment System (ICDAS) 5 or 6].<br>**4)** The cavitated lesions must allow for direct application of SDF. | **1)** Child is allergic or has sensitivity to silver.<br>**2)** Child has hereditary generalized developmental defects of enamel (e.g., amelogenesis imperfecta and dentinogenesis imperfecta).<br>**3)** Child has severe medical problems that limit participation.<br>**4)** Child requires immediate rehabilitation under general anesthesia because of severe infection or pain.<br>**5)** Antibiotic use within the last 2 weeks.<br>**6)** Any teeth that meet PUFA criteria (pulpal involvement, ulceration due to trauma, fistula, abscess). The child may still qualify if other eligible teeth with caries do not meet PUFA criteria. |

**Supplementary Table 2**. Study Stages.

<table>
<tr><td rowspan="2">Recruitment & Randomization</td><td colspan="3">• Recruitment<br>• Informed consent<br>• Randomization into one of three SDF treatment regimens (n = 15 per group) using random envelopes</td></tr>
<tr><td>Regimen 4M<br><br>WRHA Oral Health Program protocol<br><br>2 applications<br>4 months apart</td><td>Regimen 6M<br><br>Suggested by ADA<br><br>2 applications<br>6 months apart</td><td>Regimen 1M<br><br>Suggested by AAPD<br><br>2 applications<br>1 month apart</td></tr>
<tr><td>First Study Visit (V1)</td><td>• Baseline questionnaire<br>• Dental assessment<br>• Dental plaque sample<br>• First SDF + NaF varnish application</td><td>• Baseline questionnaire<br>• Dental assessment<br>• Dental plaque sample<br>• First SDF + NaF varnish application</td><td>• Baseline questionnaire<br>• Dental assessment<br>• Dental plaque sample<br>• First SDF + NaF varnish application</td></tr>
<tr><td>Second Study Visit (V2)</td><td>4 months after V1<br>• Follow-up visit questionnaire<br>• Dental assessment<br>• Dental plaque sample<br>• Second SDF + NaF varnish application</td><td>6 months after V1<br>• Follow-up visit questionnaire<br>• Dental assessment<br>• Dental plaque sample<br>• Second SDF + NaF varnish application</td><td>1 month after V1<br>• Follow-up visit questionnaire<br>• Dental assessment<br>• Dental plaque sample<br>• Second SDF + NaF varnish application</td></tr>
<tr><td>Third Study Visit (V3)</td><td>4 months after V2<br>• Follow-up visit questionnaire<br>• Dental assessment<br>• Dental plaque sample</td><td>6 months after V2<br>• Follow-up visit questionnaire<br>• Dental assessment<br>• Dental plaque sample</td><td>1 month after V2<br>• Follow-up visit questionnaire<br>• Dental assessment<br>• Dental plaque sample</td></tr>
</table>

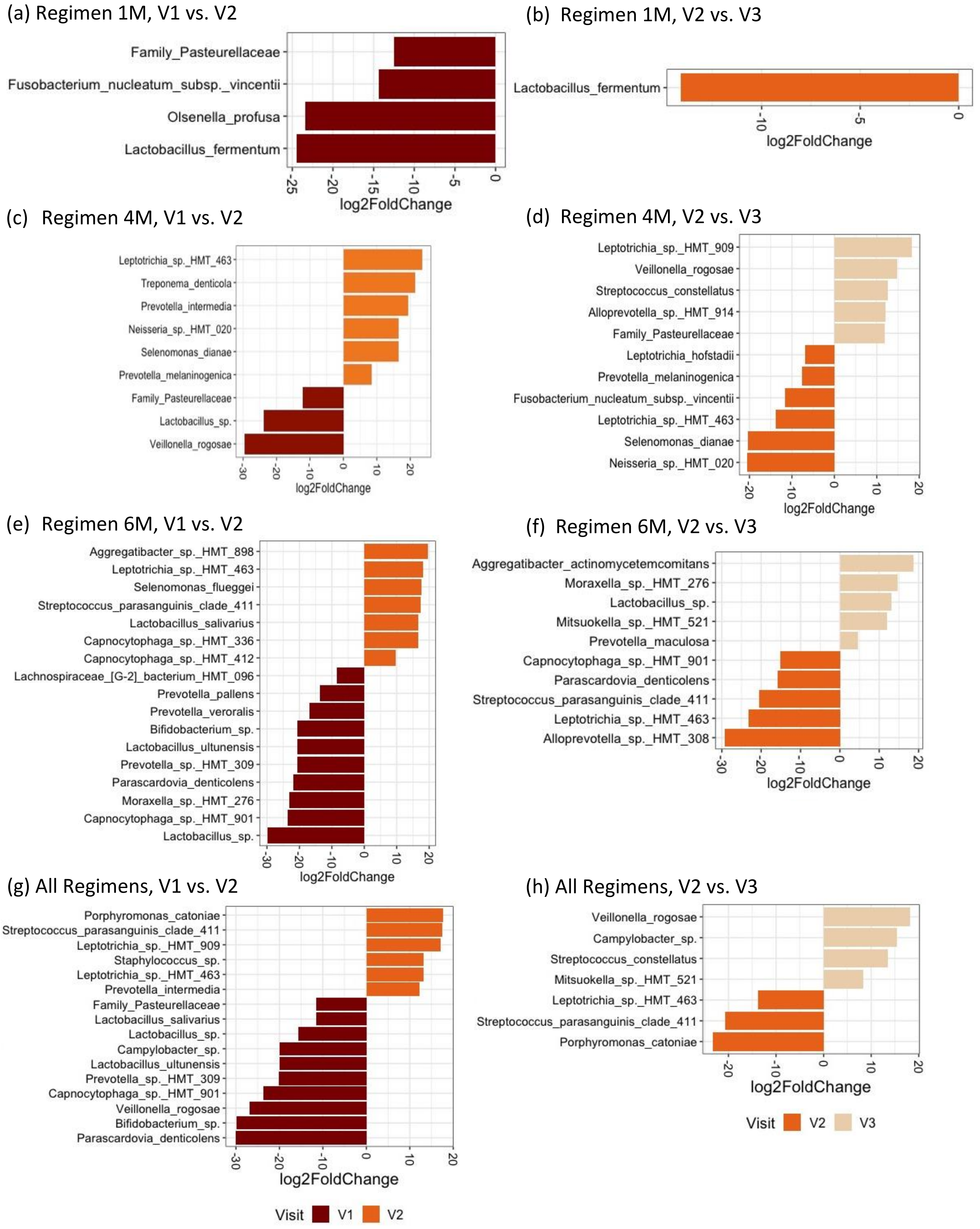
(a) Regimen 1M, V1 vs. V2
Family_Pasteurellaceae
Fusobacterium_nucleatum_subsp._vincentii
Olsenella_profusa
Lactobacillus_fermentum
log2FoldChange
(b) Regimen 1M, V2 vs. V3
Lactobacillus_fermentum
log2FoldChange
(c) Regimen 4M, V1 vs. V2
Leptotrichia_sp._HMT_463
Treponema_denticola
Prevotella_intermedia
Neisseria_sp._HMT_020
Selenomonas_dianae
Prevotella_melaninogenica
Family_Pasteurellaceae
Lactobacillus_sp.
Veillonella_rogosae
log2FoldChange
(d) Regimen 4M, V2 vs. V3
Leptotrichia_sp._HMT_909
Veillonella_rogosae
Streptococcus_constellatus
Alloprevotella_sp._HMT_914
Family_Pasteurellaceae
Leptotrichia_hofstadii
Prevotella_melaninogenica
Fusobacterium_nucleatum_subsp._vincentii
Leptotrichia_sp._HMT_463
Selenomonas_dianae
Neisseria_sp._HMT_020
log2FoldChange
(e) Regimen 6M, V1 vs. V2
Aggregatibacter_sp._HMT_898
Leptotrichia_sp._HMT_463
Selenomonas_flueggei
Streptococcus_parasanguinis_clade_411
Lactobacillus_salivarius
Capnocytophaga_sp._HMT_336
Capnocytophaga_sp._HMT_412
Lachnospiraceae_[G-2]_bacterium_HMT_096
Prevotella_pallens
Prevotella_veroralis
Bifidobacterium_sp.
Lactobacillus_ultunensis
Prevotella_sp._HMT_309
Parascardovia_denticolens
Moraxella_sp._HMT_276
Capnocytophaga_sp._HMT_901
Lactobacillus_sp.
log2FoldChange
(f) Regimen 6M, V2 vs. V3
Aggregatibacter_actinomycetemcomitans
Moraxella_sp._HMT_276
Lactobacillus_sp.
Mitsuokella_sp._HMT_521
Prevotella_maculosa
Capnocytophaga_sp._HMT_901
Parascardovia_denticolens
Streptococcus_parasanguinis_clade_411
Leptotrichia_sp._HMT_463
Alloprevotella_sp._HMT_308
log2FoldChange
(g) All Regimens, V1 vs. V2
Porphyromonas_catoniae
Streptococcus_parasanguinis_clade_411
Leptotrichia_sp._HMT_909
Staphylococcus_sp.
Leptotrichia_sp._HMT_463
Prevotella_intermedia
Family_Pasteurellaceae
Lactobacillus_salivarius
Lactobacillus_sp.
Campylobacter_sp.
Lactobacillus_ultunensis
Prevotella_sp._HMT_309
Capnocytophaga_sp._HMT_901
Veillonella_rogosae
Bifidobacterium_sp.
Parascardovia_denticolens
log2FoldChange
Visit V1 V2
(h) All Regimens, V2 vs. V3
Veillonella_rogosae
Campylobacter_sp.
Streptococcus_constellatus
Mitsuokella_sp._HMT_521
Leptotrichia_sp._HMT_463
Streptococcus_parasanguinis_clade_411
Porphyromonas_catoniae
log2FoldChange
Visit V2 V3

**Supplementary Figure 1**. Differential abundance analysis for bacterial species in Regimen (a-b) 1M, (c-d) 4M, (e-f) 6M, and (g-h) All regimens together. The figure shows the relative fold change in bacterial abundance between (a, c, e, and g) visits 1 and 2 and (b, d, f, and h) visits 2 and 3. Only bacterial taxa with FDR adjusted $p < 0.05$ are shown.

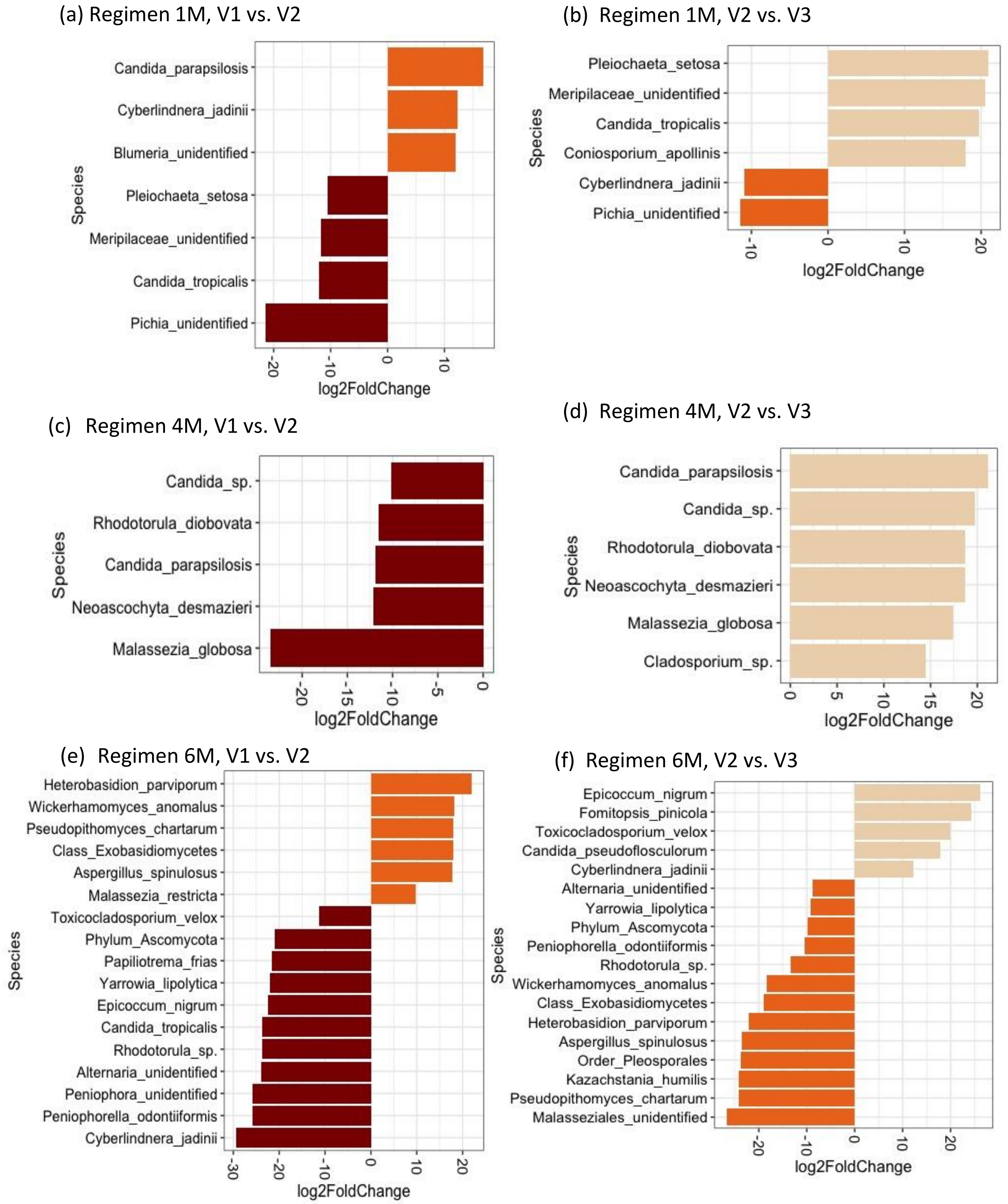


**Supplementary Figure 2**. Differential abundance analysis for fungal species in Regimen (a-b) 1M, (c-d) 4M, and (e-f) 6M. The figure shows the relative fold change in fungal abundance between (a, c, and e) visits 1 and 2 and (b, d, and f) visits 2 and 3. Only fungal taxa with FDR adjusted $p < 0.05$ are shown.

(a) All Regimens, V1 vs. V2 (b) All Regimens, V2 vs. V3

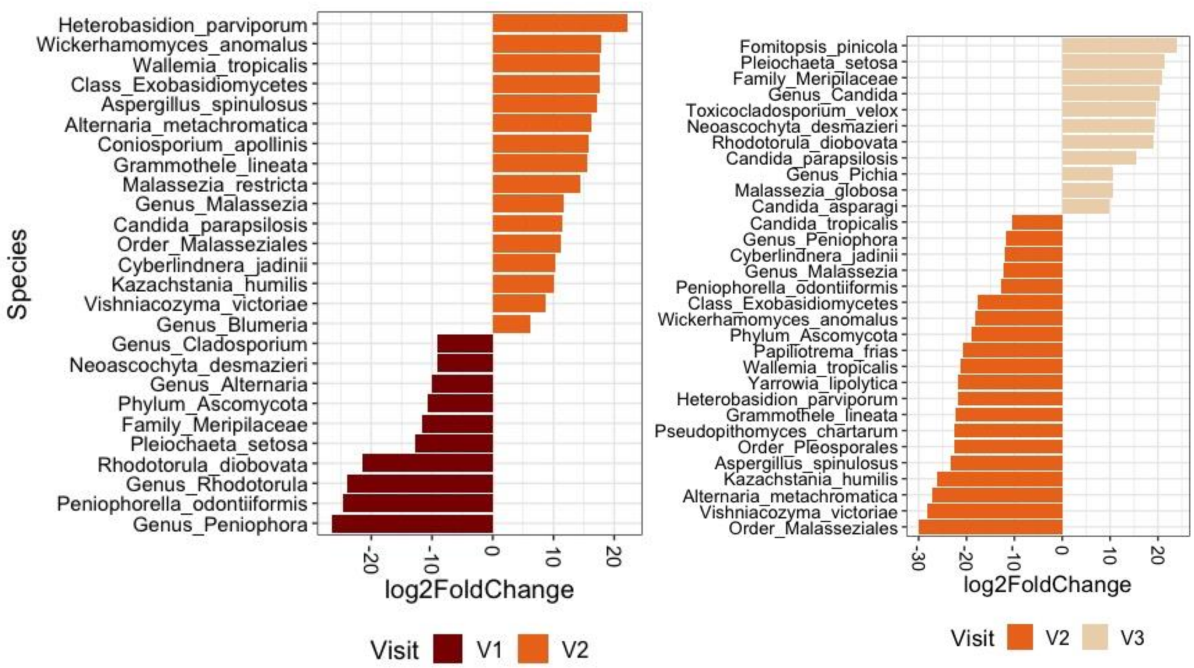


**Supplementary Figure 3**. Differential abundance analysis for fungal species in all regimens together. The figure shows the relative fold change in bacterial abundance between (a, c, e, and g) visits 1 and 2 and (b, d, f, and h) visits 2 and 3. Only fungal taxa with FDR adjusted $p < 0.05$ are shown.
